# Opportunities for Vein-to-Vein Datasets from a Blood Establishment Perspective: towards a “Pan-European Transfusion Research InfrAstructure” (PETRA)

**DOI:** 10.64898/2026.03.24.26348611

**Authors:** Sophie M. T. Wehrens, Mikko Arvas, Suzanne F. Fustolo-Gunnink, Matea Vinković Vlah, Allison Waters, Christian Erikstrup, Louise Ørnskov Drechsler, Simon J Stanworth, Katja van den Hurk

## Abstract

iii.

**Background and Objectives:** The “Pan-European Transfusion Research InfrAstructure” (PETRA) project was established to advance the use of donor, blood product, and patient datasets in Europe, aiming to benefit both patient and donor health. Here, the initial PETRA objective was to describe the landscape of existing donor and blood establishment (BE) databases.

**Materials and Methods:** An online survey was circulated to the European Blood Alliance’s BE members. The survey collected information on the feasibility of accessing donor data, and challenges and possibilities for linking these datasets with information on the associated blood products and transfusion recipients, and donors’ own health records.

**Results:** Seventeen BEs across 16 countries completed the survey. The majority could, in principle, link their donor data to product data (13 BEs (76%)) and recipient data (10 BEs (59%)), for research purposes. However, capabilities were limited and in only 29% of the BEs was the donor to recipients’ linkage an automated process. BEs reported significant challenges to achieve full vein-to-vein linkage, including legal constraints and lack of consent (11 BEs) and resources (10-14 BEs). IT and data issues as well as lack of knowledge and training were cited as obstacles by a minority of BEs.

**Conclusion:** Whilst the survey results suggest considerable interest in developing linkages between blood donors, their products, and recipients, many challenges remain due to a variety of obstacles. First steps in working towards a PETRA may be assistance to navigate legal frameworks as well as investing in resources and quality and harmonisation of data collections.

**Highlights:**

1. 17 blood establishments (BEs) in 16 countries responded to a survey on obstacles and opportunities for achieving vein-to-vein datasets.
2. In 59% of the BEs donor-to-recipient links can be established for research improving transfusion outcomes, but only in 29% this is an automated process.
3. In order to work towards a “Pan-European Transfusion Research InfrAstructure” (PETRA), legal frameworks, adequate donor consent and (financial and human) resources are the most common obstacles that require addressing.

## Introduction

Blood Establishments (BEs) and hospitals across Europe hold significant amounts of data on donors, blood products, and patients receiving these products. Better access to, and integrated analysis of, these data could lead to substantial advancements in transfusion medicine and donor health management. In some countries these datasets are both well-populated and well-integrated into the healthcare infrastructure, such as in the Scandinavian Donation and Transfusion (SCANDAT) database [1]. However, in most countries and regions, donor and patient transfusion-related datasets are scattered, and many do not link the donor to the product and the recipient for benchmarking or research purposes. ‘Vein-to-vein’ datasets are areas of intense research interest internationally as shown in the Recipient Epidemiology and Donor Evaluation Study (REDS) programme [2, 3] and roadmaps for donation and transfusion databases in Canada (CANDAT) [4] and Australia [5].

Therefore, we established a European collaborative project to work towards a “Pan-European Transfusion Research InfrAstructure” (PETRA), aiming to develop a strategic plan for further alignment of existing databases, methodologies, and research efforts. The aim of this first step towards PETRA was to describe existing databases on donors across European countries, including possibilities and barriers for accessing these data and for establishing the “vein-to-vein” link between donor, product, and the recipient.

## Methods

To build an overview of donor databases, an online survey was designed by an international interdisciplinary working group including specialists in donor and transfusion medicine research. The survey was created in Qualtrics XM, keeping in mind the heterogeneous settings of the BEs and possible responses they may (be able to) provide. Questions were made compulsory in order to achieve maximum sample size and representativeness for all questions.

In brief, the survey asked BEs about the feasibility of accessing donor data and the challenges and possibilities for linking these data to information on blood products and their recipients, as well as to the donors’ own health records. A full copy of the survey is available in the supplements. Where BEs were asked about linking donor data to transfusion recipient data, we interpreted this as the vein-to-vein (v-v) link and refer to these BEs as v-v BEs. Some questions (obstacles and plans to link to clinical databases) were asked separately in the setting of transfusion research on one hand and donor health research on the other hand as the databases, linkages and obstacles were expected to be different.

The survey was pre-tested and piloted within the working group and by working group colleagues who provided feedback on question clarity, response options, and survey flow. The survey was then circulated by the European Blood Alliance (EBA) in May 2024 to its 29 member countries via email (a list of countries is available in the supplements). BEs received this email either directly from the EBA or through intermediate bodies or committees, especially for the countries where blood services operate regionally. BEs were reminded by the EBA and working group to complete the survey and the survey was closed from October 2024. Answers that were clearly incorrect were checked with the survey respondent and corrected. All respondents gave consent for their BE’s data to be included in the inventory.

## Results

The survey was completed by respondents from 17 BEs in 16 countries. They held data of on average 259,130 donors (total 4,405,209; median 62,000; range 12,188 – 1,409,411) making an average of 775,568 donations (total 13,184,648; median 98,901; range 15,829 – 7,455,040, a mix of donation types) in the year 2023. This number of donations comprised approximately 80% of the successful donations made in the 29 EBA member countries. Eight BEs operate at a national level only and (apart from one) account for (nearly) all of the national donor information. Three organisations cover only a small percentage (1-10%) of the national number of donations, reflected in the fact that 2 BEs are active at regional/local level and 1 at regional/local as well as national level.

Most organisations agreed that linking different steps in the donor-product-recipient chain is possible; i.e. donor data can be linked for research purposes to product data by 13 establishments (76%) and to recipient data by 10 establishments (59%), investigating areas such as improving product quality and transfusion outcomes (Figure 1 and 2). However, the majority indicated that their organisation does not have fully automated processes in place to achieve this. Lack of automated processes was particularly the case for the steps that involve patients and linking their data to products and donors to improve research into transfusion outcomes.

**Figure 1.**
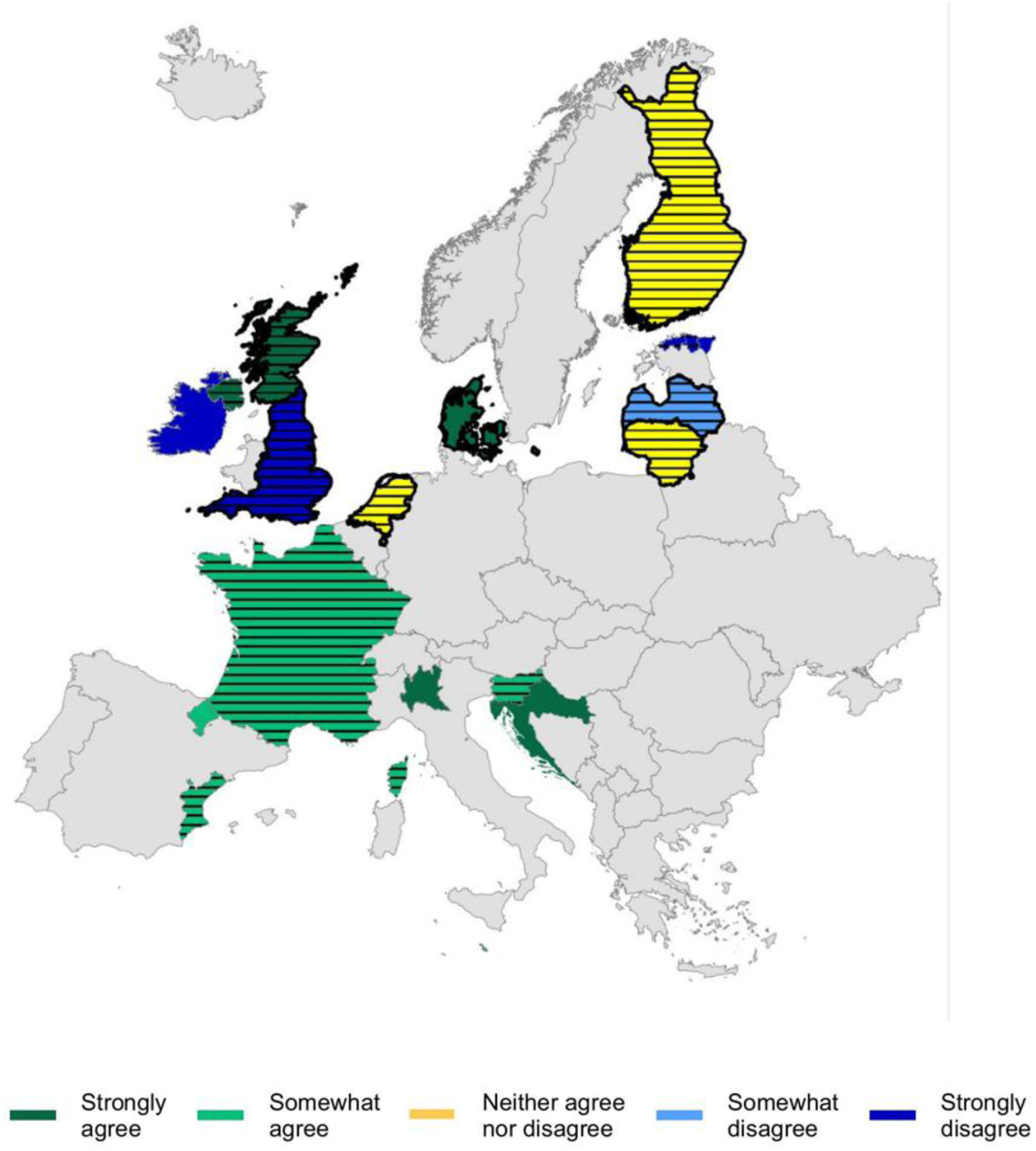
Map of Europe summarising the linkage possibilities for the responding BEs. Countries with a black outline indicated they represent their whole country and have no (additional) regional/local cover. Colours indicate to what extent respondents agreed with that statement that linking donors to recipients for research purposes is possible: strongly agree (dark green), somewhat (light green), neither agree nor disagree (yellow), somewhat disagree (light blue) and strongly disagree (dark blue). Horizontal lines indicate countries/regions that had no fully automated process for linking donors and recipients (somewhat or strongly disagreed with the statement on fully automated linkage).

**Figure 2.**
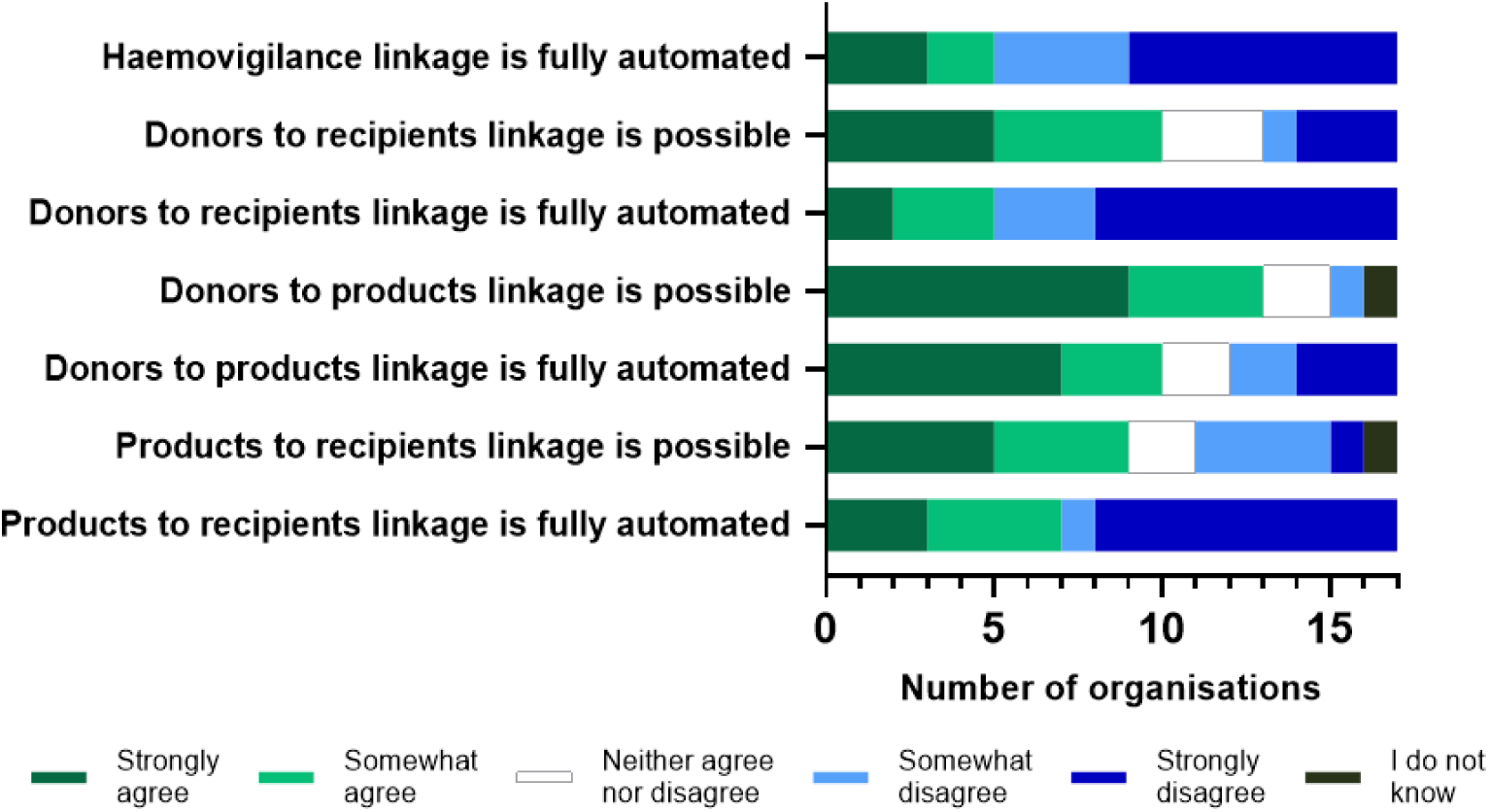
Extent to which BEs agreed on seven statements asking whether different links in the vein-to-vein chain are currently possible with their database and whether this linkage is automated. The first statement refers to linking for legal haemovigilance purposes, the other six statements ask about enabling linkages for research purposes, for example into improving transfusion outcome and product quality.

Of the 17 BEs, 65% responded with ‘yes’ or ‘maybe’ when they were asked whether they currently have the ability or have any plans to link their donor / donation database to any existing or future clinical database (Figure 3). Six BEs answered this question on linkage with transfused patients with ‘yes’ and 5 with ‘maybe’ (Figure 3A). Similarly, 4 BEs (plan to) have and 7 maybe have the ability to link their donor / donation database to the donors’ own health records for research into donor health (Figure 3B).

**Figure 3.**
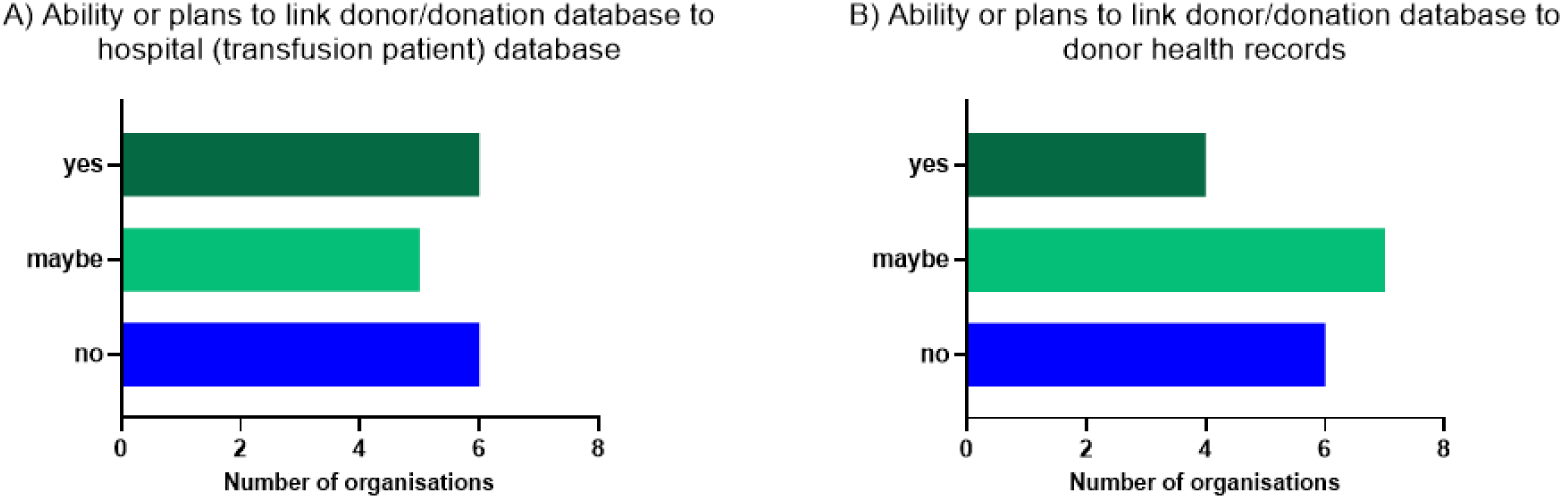
Number of BEs that have the ability or have any plans to link their donor/donation database to any existing or future patient or hospital database in order to assess transfusion outcomes (A) or existing or future (hospital) database that holds the donor’s own health records in order to follow up or research donor health (B).

Eleven (65%) of all BEs reported that legal constraints and lack of consent were extremely or somewhat likely obstacles to achieve vein-to-vein linkages to facilitate transfusion or donor health research (Figure 4A/B). Five (50%) of the 10 BEs that reported they could link donor to recipient data (v-v BEs) stated legal constraints and lack of consent as likely obstacles (data not shown). Lack of staff or financial resources was cited as a likely obstacle for 14 (82%) of all BEs or 7 of the v-v BEs when using these databases to support transfusion research and by 10 (59%) of all BEs or 4 of the v-v BEs to facilitate donor health research. Difficulty in finding variables in or matching between datasets was indicated by 9 BEs (53%), 4 of the v-v BEs, as a likely obstacle for donor health research and lack of compliance in completing records by 9 organisations (53%), 3 of the v-v BEs, for transfusion research. IT issues, lack of knowledge and training, and the absence of a unique identifier were cited as specific research obstacles in a minority (less than 7) of blood establishments.

**Figure 4.**
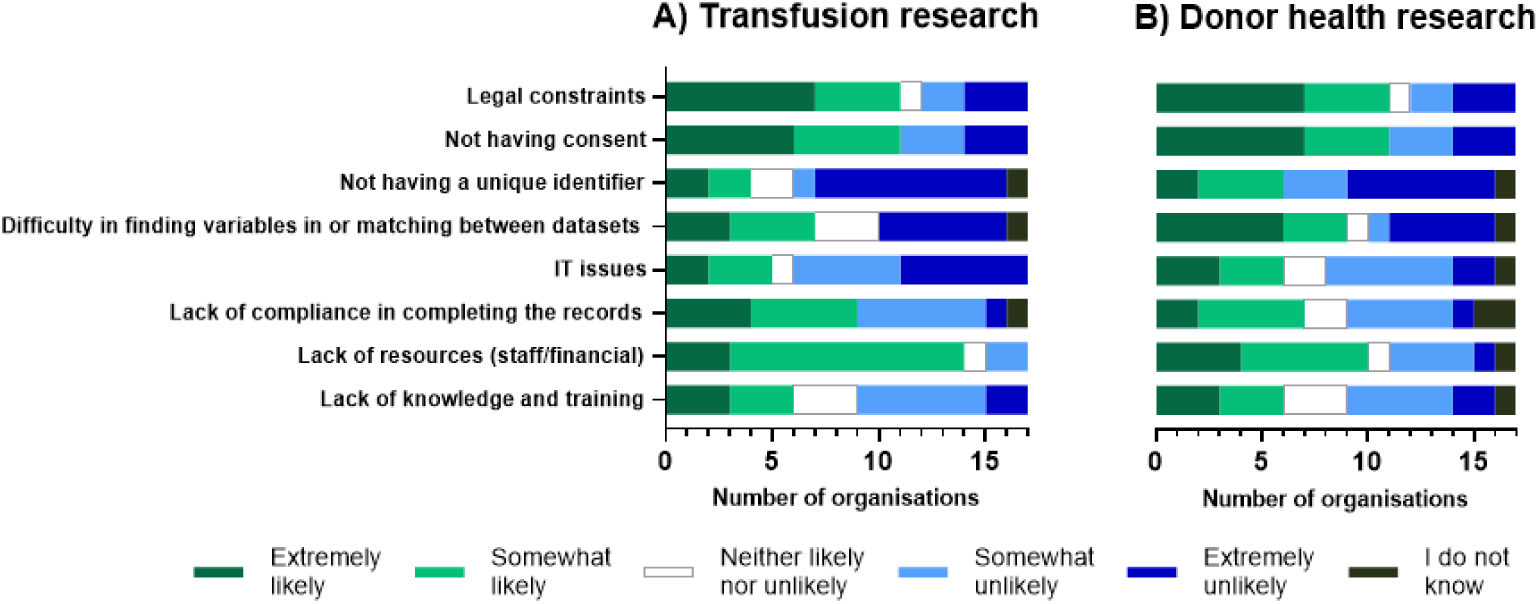
Likelihood that BEs will encounter or observe eight different issues or obstacles when linking or trying to link donor, product, and patient data for research into transfusion outcomes (A) or donor health (B). For some of the statements organisations were provided with examples (see supplement).

## Discussion

This study demonstrated substantial variability in the capabilities of 17 BEs in 16 European countries to access and connect donor, product, and recipient data. Only 59% of BE’s were (in principle) able to establish v-v linkages, and just 29% could do this in a fully automated way. The results provide information on barriers for the creation of vein-to-vein infrastructures that would allow for the conduct of high-quality and efficient clinical transfusion and donor health research, as well as optimal quality control and stock management for BEs and hospitals.

Obstacles for further establishing linkages were mainly legal constraints not having donor consent and lack of financial and personnel resources Interestingly, in those BEs that were able to establish the link between donors and recipients, obstacles were still observed and the trend was similar in terms of the percentage of BEs that reported the obstacles for the two different purposes (transfusion and donor health research). As BEs were asked if they would encounter *or* observe these issues/obstacles when linking *or* trying to link data, it may imply that both groups of BEs recognised these obstacles in a similar way but that those who had successfully established the link, had overcome these obstacles. In order to keep the survey short, no questions were added to assess the effort it took BEs to establish the linkages. However, ongoing conversations indicate that many organisations need to spend considerable time and resources on linking, even though they are (ultimately) able to establish one. In line with our findings, other roadmaps to clinical and vein-to-vein datasets have also suggested that challenges in the ethical-legal, funding and data quality areas as well as other barriers need to be overcome to reach successful implementation [4, 6, 7]. Once established, however, the potential of such datasets for advancing clinical and research priorities is evident [1–3, 7, 8]. Those integrated datasets enable and improve, for example, clinical practice such as clinical audit and feedback [6], surveillance for unknown transfusion-transmitted pathogens [9] or studies into donor and component factors affecting recipient outcomes [10–12].

Contrary to the expectation that the different setting for transfusion compared to donor health research would result in different obstacles, observed barriers were similar in both settings, although lack of resources was more frequently reported to hinder linking for transfusion than for donor health research. On the other hand, plans to link donor/donation databases to clinical databases may be slightly more concrete in the field of transfusion than donor health research.

There are strengths and limitations to our survey. A consideration in the interpretation of the results is the geographical spread and heterogeneity (e.g. in national versus regional organisation of BEs) in respondents. Although we did not systematically invite BEs to respond to the survey this spread may have resulted in a good degree of representation. On the other hand, heterogeneity also made it difficult to design a survey that would fit all BEs. For example, a more precise estimate of the extent to which the BEs were able to establish linkages and which specific parts are automated (if not done fully) cannot be derived from the current questions and may have been interpreted differently by different BEs.

Our findings suggest that investment in resources as well as assistance to navigate legal frameworks (including possibilities and exemptions under the General Data Protection Regulation (GDPR) and adequate (modular) informed consent may improve the potential to link these existing databases. Nearly all BEs were interested in further investigating possibilities to map and improve integration of donor, donation and patient data for transfusion and donor health research. Some BEs have already made changes following completion of the survey.

The findings of this survey will enable collaborative development of a strategic plan for a Pan-European transfusion infrastructure Work to progress these areas will involve standards and recommendations in line with European regulations such as the GDPR, European Health Data Space (EHDS), Substances of Human origin (SoHo) and guidelines from the European Directorate for the Quality of Medicines & HealthCare (EDQM). Ultimately, these infrastructures will enable further research aimed at better donor health management, monitoring blood component utilisation and improving patient outcomes.

## Sources of research support

Funding for this project has been provided in part through an Agreement with the European Blood Alliance (EBA). The contents of this document do not necessarily reflect the views and policies of the EBA, nor does mention of trade names or commercial products constitute endorsement or recommendation of use.

## Data availability statement

Data related to this publication are held at Sanquin Blood Supply Foundation. Requests to access these data in line with relevant regulations and agreements may be made to the corresponding authors.

## ii. Acknowledgements

## Author contributions

SW, MA, SF, AW, CE, SS and KvdH initially conceived the idea; SW, MA, SF, MV, AW, CE, LOD, SS and KvdH designed the survey; SW analysed the survey; SW, MA, SF, MV, AW, CE, LOD, SS and KvdH wrote, reviewed and edited the manuscript; all authors read and approved the final manuscript.

We thank Professor Emanuele Di Angelantonio for sharing his experience and insights in the conceptualisation phase of the project. We are also grateful to all the individuals of the participating organisations that took the time to carefully complete the survey:

Banco de Sangre y Tejidos de Navarra - Hospital Universitario de Navarra (Spain)

Centro de Transfusion de la Comunidad Valenciana (Spain)

Croatian Institute of Transfusion Medicine (Croatia)

The Danish Blood Centers (Denmark)

Finnish Red Cross Blood Service (Finland)

Fondazione Ca’ Granda IRCCS Ospedale Maggiore Policlinico Milano (Italy)

French Blood Establishment (France)

Irish Blood Transfusion Service (Ireland)

Malta National Blood Transfusion Service and Mater Dei Hospital Blood Bank (Malta)

National Health Service Blood and Transplant (England)

The Northern Ireland Blood Transfusion Service (Northern Ireland)

North Estonia Medical Centre Foundation (Estonia)

Public Institution National Blood Centre of Lithuania (Lithuania)

Sanquin Blood Supply Foundation (The Netherlands)

Scottish National Blood Transfusion Service (Scotland)

Slovenian Institute for Transfusion Medicine (Slovenia)

State Blood Donor Centre (Latvia)

## Data Availability

Requests to access the data related to this publication may be made to the corresponding authors in line with relevant regulations and agreements.

## ix. Appendices

N/A

Member countries of the European Blood Alliance receiving the survey in May 2024, listed in alphabetical order

1. Austria
2. Belgium
3. Croatia
4. Denmark
5. Estonia
6. Finland
7. France
8. Germany
9. Greece
10. Iceland
11. Ireland
12. Italy
13. Latvia
14. Lithuania
15. Luxembourg
16. Malta
17. Netherlands
18. North Macedonia
19. Norway
20. Portugal
21. Serbia
22. Slovenia
23. Spain
24. Sweden
25. Switzerland
26. UK - England
27. UK - Northern Ireland
28. UK - Scotland
29. UK - Wales

## PETRA - donor databases

Start of Block: Intro

Q1.1 Dear colleague,

Welcome to this survey towards a **Pan-European Transfusion Research infrAstructure (PETRA),** a project funded by the European Blood Alliance, led by Sanquin Blood Supply Foundation. We would be very grateful for your support and responses to this short survey aimed at understanding the capabilities of donor databases.

The application of data science in health research offers significant opportunities in transfusion medicine. There are also opportunities with better linkage of donor and patient data and alignment of infrastructure across Europe. The overarching aim of this PETRA project is to inform a strategic plan to direct further research at the European level, including aligned ’vein-to-vein’ research infrastructures.

In this survey, we are also are seeking your interest in joining our project group to explore the feasibility of electronic data collection to efficiently understand ‘where blood goes’, and to investigate a clinical research question related to a select number of patient groups that are potentially over- or under-transfused.

The aim of this survey is to obtain an overview of the existing electronic platforms and datasets used in the blood supply chains across Europe. The survey will collect background and meta-data on existing donor- and product datasets. In a later survey we will approach organisations that hold clinical or patient datasets that might be relevant to blood transfusion. The results will be used to produce an inventory of European blood donor-, product- and transfusion databases as a basis for the strategic plan for a pan-European research infrastructure.

- Please note that given the nature of the survey, there may be a need to collect answers on certain parts of the survey from other individuals in your organisation or different colleagues. Rather than forwarding the survey for others to add to, we recommend that one person collects all answers and keeps track of survey completion. Even partially completed, your answers will be saved, so you won’t have to take the survey from the beginning all over again if you need to pause. If you wish to do this, please make sure you use the same internet browser on the same device.
- If you wish to download a PDF-ready-to-print-version of this survey before completing it online, you can click on <u>the link to the PETRA survey</u>. You will also be able to view and print an overview of your responses before submitting them.
- We will process any personal data you provide in line with <u>Sanquin’s Privacy Statement</u>. By completing this survey you consent for metadata about your dataset and organisation to be mentioned in the inventory. We will only collect aggregated data about donors, products and patients and therefore donors and patients will never be identified in outcomes of the PETRA inventory, reports and publications.
- We estimate that filling out the survey takes around 30 minutes, depending on whether you have to collate the information, and you have up to 26th of May 2024 to complete this survey.

If you have any questions beforehand or while you are filling out the survey, please contact the PETRA team via e-mail at.

**Thanks in advance for your time and effort!**

On behalf of the PETRA team,

Dr. Sophie Wehrens

*Dr Katja van den Hurk, Dr Suzanne Fustolo-Gunnink and Dr Sophie Wehrens (Sanquin Blood Supply Foundation, The Netherlands)*

*Prof Christian Erikstrup and Louise Ørnskov Drechsler (Aarhus University Hospital, Denmark) Matea Vinković (Croatian Institute of Transfusion Medicine, Croatia)*

*Prof Simon Stanworth (European Haematology Association Specialised Working Group in Transfusion, the NIHR Blood and Transplant Unit in data, Oxford, UK)*

*Dr Mikko Arvas (Finnish Red Cross Blood Service, Finland)*

*Dr Allison Waters and Eileen Ryan (Irish Blood Transfusion Service, Children’s Health Ireland, Crumlin Hospital, Ireland)*

*Prof Emanuele Di Angelantonio (NHSBT, University of Cambridge, UK)*

Q2.1 **Do you consent for your organisation’s name and website to be listed in the PETRA inventory?**

*Please note that if you select ’no’, you can no longer complete the survey.*

- Yes
- No

__________________________________________

Q3.1 **Do you consent for your name and contact details to be listed in the PETRA inventory?**

*Your organisation will be mentioned in the inventory by default.*

- Yes
- No

__________________________________________

You can now start completion of the survey. For your information, you can still access <u>the link</u> <u>to the survey pdf</u> whilst completing it; it includes the instructions as well as all the questions.

You will also be able to (re)view and print your responses before submitting them.

__________________________________________

Q4.1 **What is your name?**

*i.e. the person providing the information for this survey and whom we may contact if we have any questions about your replies.*

__________________________________________

**Q4.2 What is your e-mail address?**

*We will only use this e-mail address to ask for clarification on the answers provided (if necessary) and to share the results with you.*

__________________________________________

Q5.1 **What is the name of the primary organisation that you represent?**

__________________________________________

Q5.2 **Which of the descriptions below best describes your organisation?**

*select all that apply*

׈ Blood collection service
׈ Blood product processing and release
׈ University/Academic hospital
׈ General hospital
׈ University
׈ Research (Institute)
׈ Other, please specify

__________________________________________

Q5.3 **What is your organisation’s website?**

__________________________________________

Q5.4 **In what country is your organisation active?**

*Please choose from the auto-complete list (country names are in English)*

__________________________________________

Q5.5 **Is your organisation active at national or regional level?**

- National
- Regional/local
- Both national and regional/local

__________________________________________

Q6.1 **The following questions are about the number of donors or donations your organisation holds information on. Please answer all questions; use whole numbers without decimal places only, enter ’0’ if your organisation does not hold this information or ’I do not know’ if you do not know the percentage. Where we ask for the percentage we mean the number of donors or donations your organisation holds information on as a percentage of the national number of donors or donations; please indicate if you had to estimate this number. Please provide the numbers for the year 2023.**

Q6.2 **How many donors does your organisation hold information on?**

*Please include all donors that donate blood (products) for transfusion purposes. Please complete both fields below and provide the total number of individuals that made a donation in the year 2023.*

׈ absolute number in the organisation (please use numbers only; enter 0 if your organisation does not hold donor information)

__________________________________________

׈ percentage of total national number of donors who donated in 2023 (please use numbers only; enter 0 if your organisation does not hold donor information. Or ’I do not know’ if you do not know this information)

__________________________________________

׈ I/we estimated the percentage of the total national number above (please select if you estimated rather than know or calculated the exact percentage)

__________________________________________

Q6.3 **How many donations does your organisation hold information on?**

*Please complete both fields below and provide the total number of donations in the year 2023.*

׈ absolute number in the organisation (please use numbers only; enter 0 if your organisation does not hold product/component information)

__________________________________________

׈ percentage of total national number of donations (please use numbers only; enter 0 if your organisation does not hold product/component information. Or ’I do not know’ if you do not know this information)

__________________________________________

׈ I/we estimated the percentage of the total national number above (please select if you *estimated* rather than *know* or *calculated* the exact percentage)

Q6.4 Please explain how you have defined a donation for this survey:

- We have included all succesful donations
- We have included all donation attempts
- We have used another defenition / selection criteria, namely:

Q6.5 Are there any particulars that we should know about the selection or inclusion criteria that you used to provide the numbers of donors and/or donations above in order to interpret these numbers?

__________________________________________

__________________________________________

__________________________________________

__________________________________________

__________________________________________

Q7.1 **The following questions assess electronic systems and implementation levels at your organisation.**

__________________________________________

Q7.2 Who is your electronic donor / blood bank management provider for your donor / donation database (for example eProgesa)?

׈ Our provider is (please specify software and current version in place):

__________________________________________

׈ ⊗Our organisation does not have an electronic donor / blood bank management provider
׈ ⊗I do not know
׈ ⊗Our organisation uses manual processes in addition to an electronic blood bank system

Q7.3 Who is your electronic Laboratory Information Management System (LIMS) provider?

׈ Our provider is (please specify software and current version in place):

__________________________________________

׈ ⊗Our organisation does not have a LIMS
׈ ⊗Data are fed back entirely into our donor database
׈ Data are partly fed back into our database, please describe:

__________________________________________

׈ Data are not fed back into our donor database
׈ ⊗I do not know
׈ Our organisation uses manual processes in addition to an electronic laboratory system

Q8.1 **The following questions aim to get an overview of the linkage and access possibilities that currently exist for donor/donation data and samples held by your organisation and any possibilities or obstacles you experience.**

__________________________________________

Q8.2 Do you contribute to a **national** database that holds a dataset containing predefined data types (either by directly adding donor and/or product data or by letting the central database copy or view data from your organisation’s database)?

- Yes, please: (1) provide the name of the database, (2) specify whether this is a donor / product or combined database (one that also holds patient data) and (3) how often and when data accessible from this database are updated:

__________________________________________

- No

Q8.3 Please indicate to what extent you agree with the statements on your database and whether it currently supports a ’vein-to-vein’ link from donor to product to a recipient database (e.g. at hospital level).

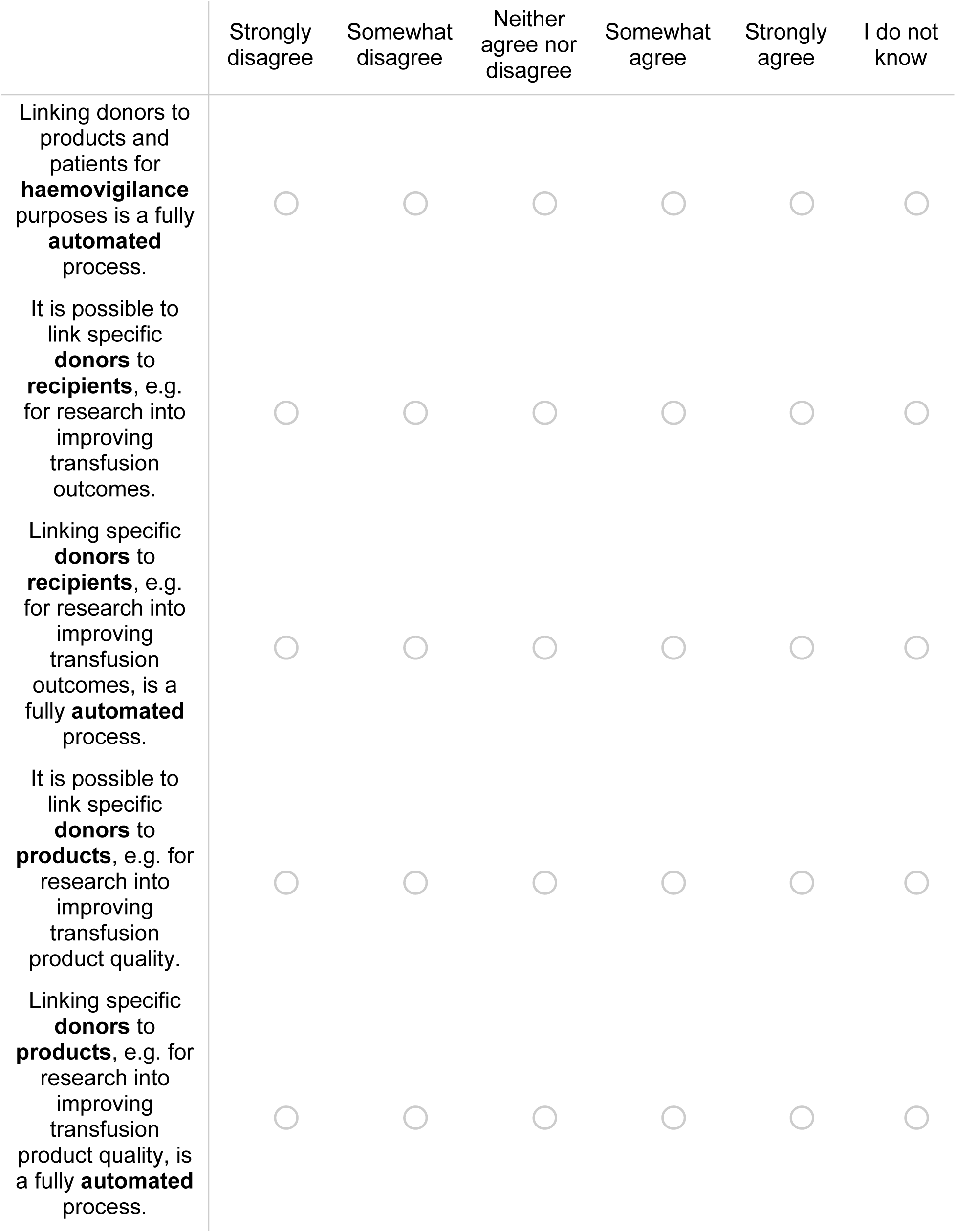
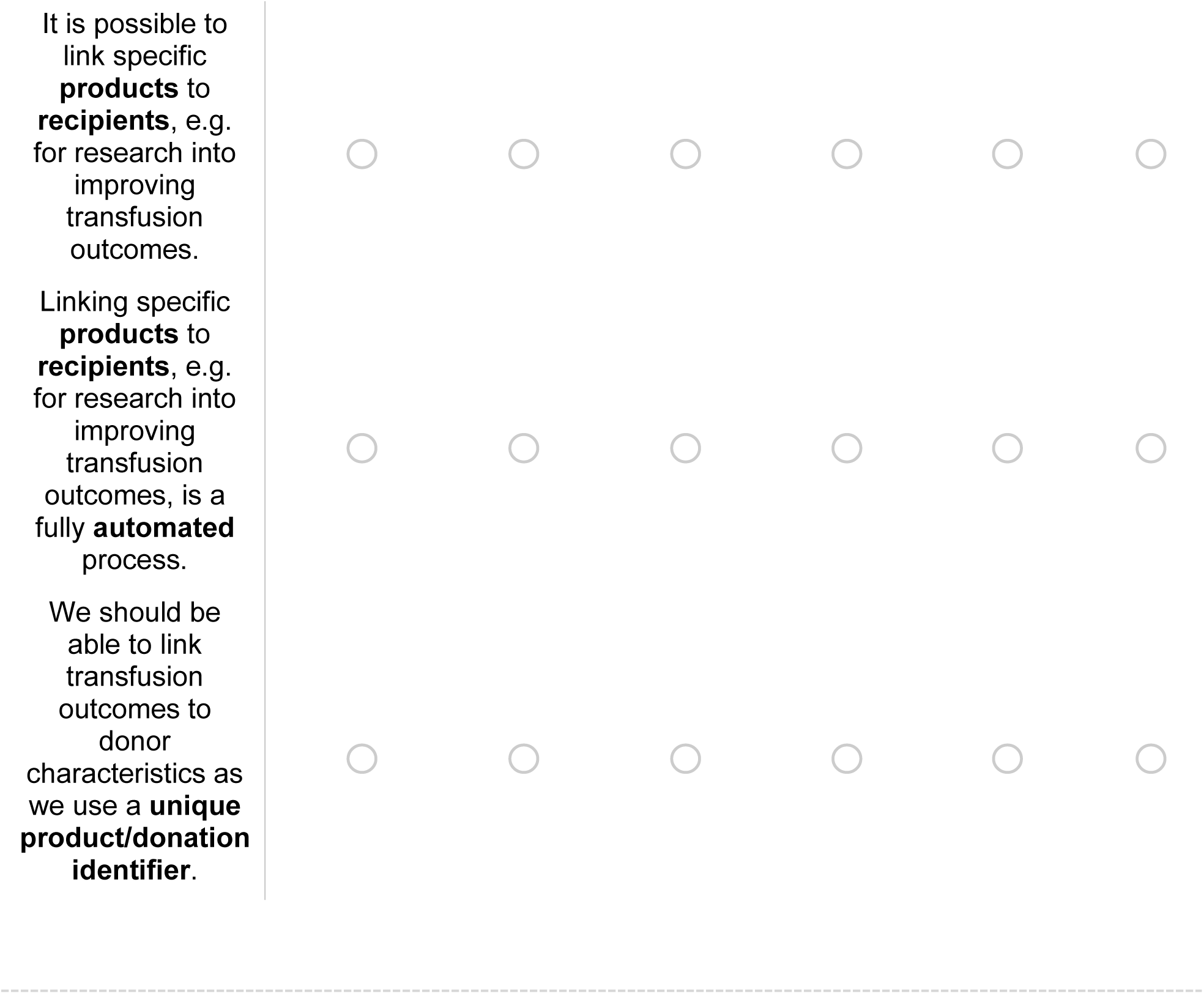

Q8.4 Can you or do you have any plans to link your donor/donation database to any existing or future *patient or hospital database* in order to assess *transfusion outcomes*?

- No
- Maybe, please provide, if possible, the name of the database, website and details of a contact person whom we may contact for further information: *Please ensure you have permission from this contact person and the owner of the database to provide us with this information* __________________________________________
- Yes, please provide, if possible, the name of the database, website and details of a contact person whom we may contact for further information: *Please ensure you have permission from this contact person and the owner of the database to provide us with this information.* __________________________________________

Q8.5 **How likely is it that you will encounter or observe the below issues/obstacles for your organisation when linking or trying to link donor, product and *patient data for research into transfusion outcomes*?**

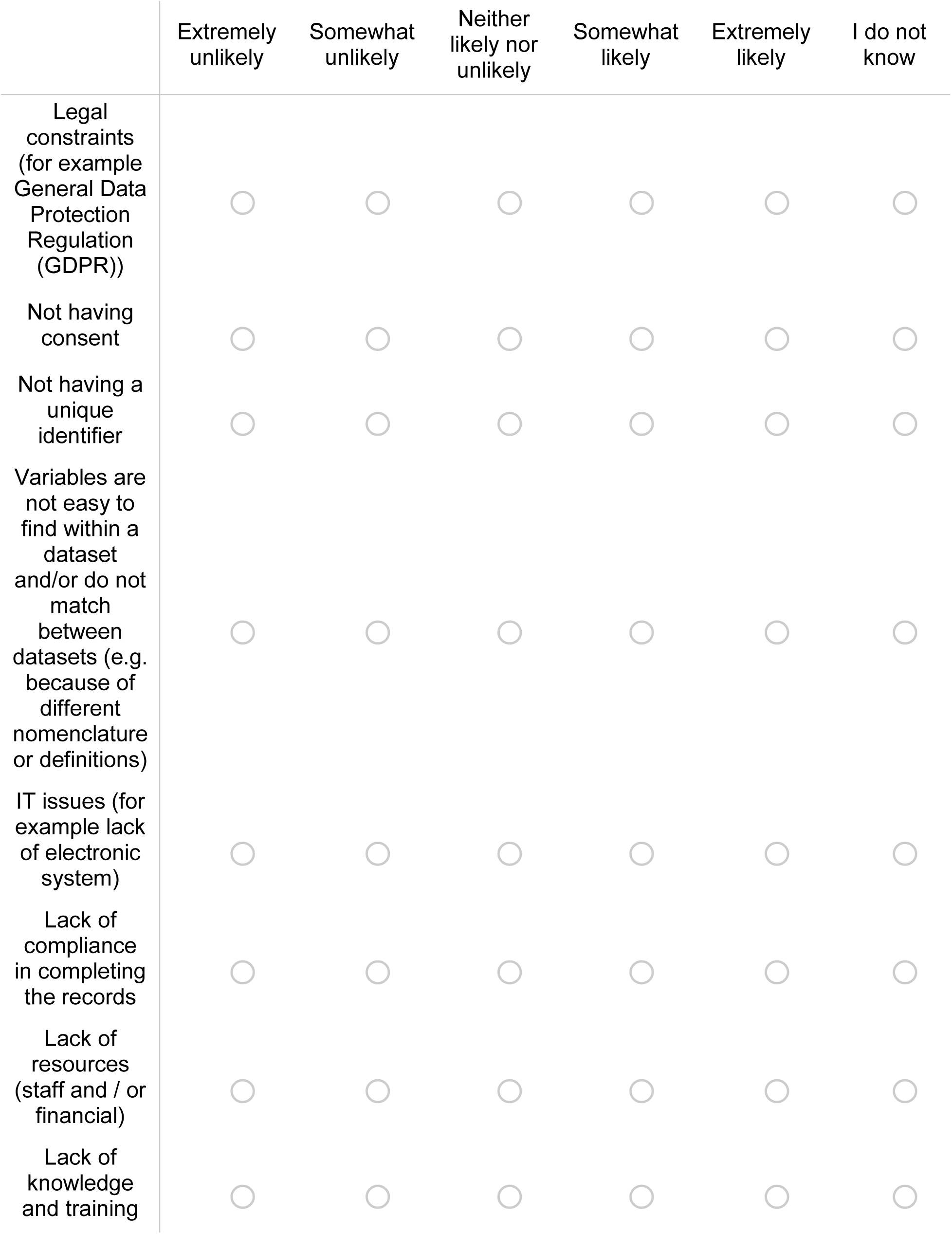
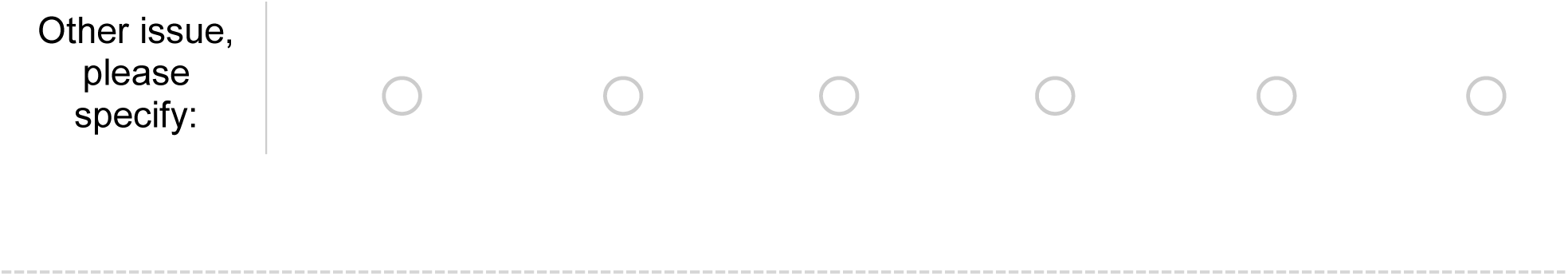

Q8.6 If you cannot currently make the link, between donor, product and patient (recipient), do you have an idea which kind of solutions could enable this linkage in theory?

- No
- Yes, please specify: __________________________________________

Q8.7 Can you or do you have any plans to link your donor/donation database to any existing or future (hospital) database *that holds the donor’s own health records* in order to follow up or research *donor health*?

- No
- Maybe, please provide, if possible, the name of the database, website and details of a contact person whom we may contact for further information: *Please ensure you have permission from this contact person and the owner of the database to provide us with this information* __________________________________________
- Yes, please provide, if possible, the name of the database, website and details of a contact person whom we may contact for further information: *Please ensure you have permission from this contact person and the owner of the database to provide us with this information.* __________________________________________

Q8.8 **How likely is it that you will encounter or observe the below issues/obstacles for your organisation when linking or trying to link donor data to *donor health records for research into donor health*?**

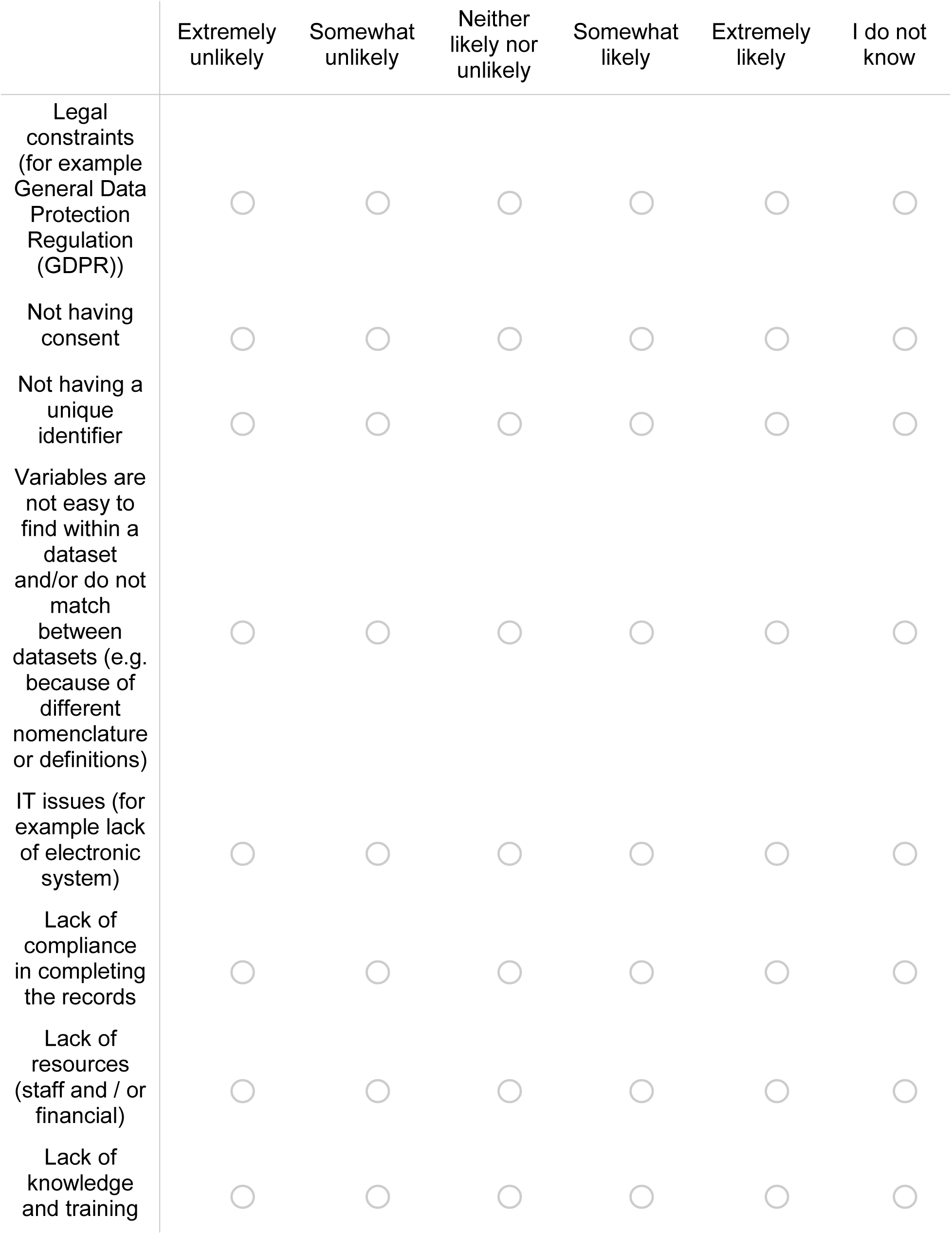
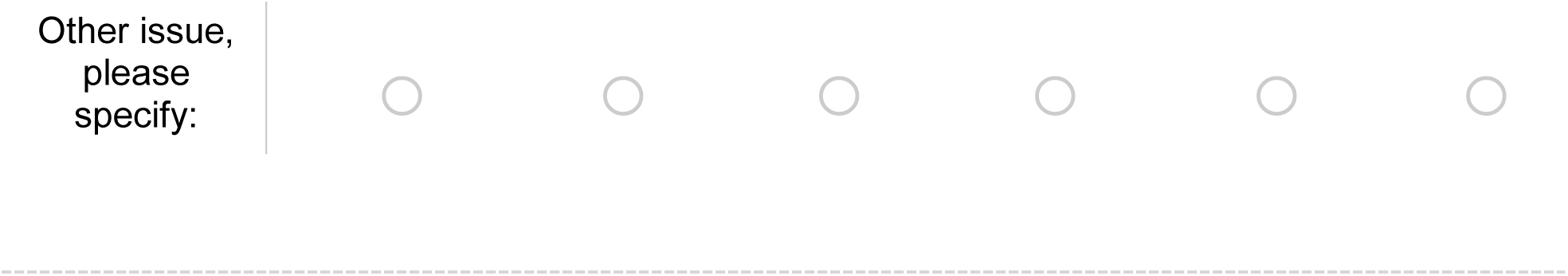

Q8.9 If you cannot currently make the link, between your donor data and donor health records, do you have an idea which kind of solutions could enable this linkage in theory?

- No
- Yes, please specify: __________________________________________

Q8.10 **How is access to data/samples held by your organisation controlled?** *Here we mean access that is required to carry out activities that are directly related to safeguarding the transfusion chain (donors, blood products and recipients)*

- Internal access for all employees
- Internal access for certain authorised departments or employees
- Internal access plus sharing with specific external partners for health care (e.g. General Practitioner, medical specialist)

Q8.11 **How is external access to data/samples held by your organisation governed arranged for (clinical) research?**

*Assuming that access for external parties for (clinical) research needs to meet certain conditions can you select all conditions that may need to be met or checked or bodies/individuals that need to be consulted in order to gain access?*

׈ Specific clinical / research department that holds the data/tissue
׈ Data/tissue access committee
׈ Internal governance approval (e.g. to check compliance and feasibility)
׈ External governance approval (e.g. to check compliance and feasibility)
׈ Internal ethical approval (e.g. Institutional Review Board, Medical Ethics Committee)
׈ External ethical approval (e.g. Institutional Review Board, Medical Ethics Committee)
׈ Assessment by an internal Data Protection Officer or Legal Counsel
׈ Other, please specify:

__________________________________________

׈ ⊗All data that are available to external parties are open access
׈ ⊗I do not know

Q9.1 **Would you be interested in being involved in our Proof of Concept study (“where does blood go?”)?**

*Here we will asses feasibility of electronic data collection to investigate a clinical research question related to a select number of patient groups that are potentially over- or under-transfused. We would work together to create a small vein-to-vein database with actual patient/donor data, to see how that could be organised and scaled up in the future. If you select ’yes’ or ’maybe’ we may contact you via the email address provided earlier in this survey.*

*Whether we contact you or not may, for example, depend on capacity and requirements for the clinical question to be investigated.*

- Yes
- Maybe
- No

Q10.1 **Is there any information on: existing electronic platforms and datasets used in the blood donation and transfusion chains possibilities or challenges for linking these datasets that you would like to provide, which did not fit in the questions above?** *If so, please explain below:*

__________________________________________

__________________________________________

__________________________________________

Q10.2 **Are there any comments you would like us to add to the inventory for your organisation?**

__________________________________________

__________________________________________

__________________________________________

__________________________________________

Q11.1 In the following questions we will ask about the data/samples that your organisation collects or holds in order to get an idea of the data types that can be linked. Please include data/samples that you include as part of the ***routine process*** during donor checks, blood collection or blood product processing/release in the majority of that group. We mean all measurements that are obtained under your current policy even if not applicable to every donor, donation, product or patient (e.g. ferritin measurement may be part of the current donor check but not be measured every donation; if this is the case, please still select this parameter).

These questions on data / samples are optional in order to save you time. If you do not wish to answer these questions, simply skip the questions by clicking the ’next’ button.

Q11.2 Which ***donor*** data do you collect / hold as part of the ***routine process****? Please select all options that apply to your organisation.*

׈ my organisation does not collect / hold donor data or samples
׈ Age
׈ Weight
׈ Sex
׈ Other demographics
׈ Blood pressure
׈ Capillary hemoglobin - primary method for donor screening
׈ Capillary hemoglobin - secondary method for donor screening in specific cases
׈ Infectious disease screening
׈ Ferritin measurements
׈ Donation and deferral history
׈ Adverse reactions to blood donation
׈ Socioeconomic data (e.g. education, job type, income)
׈ Lifestyle data (e.g. physical activity, diet, smoking)
׈ Complete full blood count data
׈ Hemoglobin only - primary method for donor screening
׈ Hemoglobin only - secondary method for donor screening in specific cases
׈ Other or specific set of full blood count paramters, please specify:

__________________________________________

׈ “Omics”, such as genomics data
׈ genetics - smaller datasets, such as genotypes or SNPs
׈ Clinical biochemistry
׈ Other laboratory measurements, please specify:

__________________________________________

׈ Diagnosis codes
׈ Prescriptions
׈ plasma
׈ serum
׈ buffy coats
׈ peripheral blood mononuclear cells (PBMCs)
׈ DNA
׈ fresh samples
׈ involvement in research
׈ re-contact for further research
׈ linkage with health records
׈ other data or samples, please specify:

__________________________________________

Q11.3 Which ***blood product and component data*** do you collect / hold as part of the ***routine process****?*

*Please select all options that apply to your organisation*.

׈ ⊗my organisation does not collect / blood product or component data
׈ type of component
׈ the DATE at which different processing steps were carried out
׈ the TIME at which different processing steps were carried out
׈ quality control data
׈ other data, please specify:

Q12.1 **You have now come to the end of the survey. On the next page you will find a summary of your responses. You can still edit these by using the left arrows to go back.**

**Once you click the right arrow (’next’ button) below the summary, your responses will be recorded and you are no longer able to go back and edit them.**

